# Prevalence of Insomnia and Its Association with Depressive Symptoms Among Patients Recovered from COVID-19 After Hospitalization: A Cross-Sectional Study in Zanjan, Iran

**DOI:** 10.64898/2026.07.26.26358944

**Authors:** Nosrat Kashefi Sis, Pourya Kashefi Sis, Ali Shendabadi

**Affiliations:** Psychiatry Department, School of Medicine, Zanjan University of Medical Sciences, Sobuti, Zanjan, Iran; School of Medicine, Hormozgan University of Medical Sciences, Emam Hossein, Bandar Abbas, Iran; College of Interdisciplinary Sciences and Technologies, University of Tehran, N. Kargar, Tehran, Iran

**Author notes:** Contributing authors.

**Keywords:** COVID-19, post-COVID insomnia, post-COVID depression

## Abstract

**Background:** COVID-19 has been associated with persistent neuropsychiatric symptoms after recovery, including sleep disturbance and depressive symptoms. Patients who required hospitalization may be particularly vulnerable to post-acute psychological complications. This study aimed to determine the prevalence of insomnia and its association with depression among patients who had recovered from COVID-19 after hospitalization.

**Methods:** This analytical cross-sectional study was conducted in 2023 at Shahid Beheshti Hospital, Zanjan, Iran. A total of 70 patients who had been hospitalized with PCR-confirmed COVID-19 and had recovered for at least 6 months were included. Data were collected from medical records and telephone interviews. Insomnia severity was assessed using the Insomnia Severity Index. Descriptive statistics, univariable and multivariable linear regression were used to analyze factors associated with insomnia severity.

**Results:** The mean ISI score was 8.06 ± 6.57. Overall, 40.0% of participants had at least subthreshold insomnia, and 17.1% had moderate to severe clinical insomnia. 30.0% of participants had some degree of depressive symptoms. In univariable analysis, higher depression score and lower educational level were associated with greater insomnia severity. In the multivariable regression model, depressive symptom severity remained independently associated with insomnia severity; each one-point increase in BDI-II score was associated with a 0.50-point increase in ISI score (adjusted *β* = 0.50; 95% CI: 0.34–0.66; p < 0.001).

**Conclusion:** Insomnia and depressive symptoms were common among patients who had recovered from COVID-19 after hospitalization. Depressive symptom severity was independently associated with greater insomnia severity. Routine screening for sleep disturbance and depressive symptoms may be beneficial in post-COVID follow-up care, particularly among previously hospitalized patients.

## 1 Introduction

Coronavirus disease 2019 (COVID-19) is an acute infectious disease caused by severe acute respiratory syndrome coronavirus 2 (SARS-CoV-2) [1]. First identified in Wuhan, China, in late 2019, the disease rapidly spread across countries and was declared a global pandemic by the World Health Organization in March 2020 [2]. COVID-19 imposed a substantial burden on health systems worldwide and also affected Iran considerably, with high numbers of infections, hospitalizations, and deaths during successive waves of the pandemic [3]. Although the clinical presentation of COVID-19 is variable, the most common manifestations include fever, cough, dyspnea, fatigue, myalgia, headache, sore throat, and loss of smell or taste; in some patients, the disease may progress to severe respiratory and systemic complications requiring hospitalization [4].

Beyond its acute physical manifestations, COVID-19 has been associated with important psychological and psychiatric consequences [5]. Both the infection itself and the broader pandemic context including fear of illness or death, social isolation, quarantine, reduced social interaction, financial stress, and uncertainty have been linked to increased emotional distress and mental health problems [6, 7]. Previous studies have reported higher rates of anxiety, depression, stress-related symptoms, sleep disturbance, and insomnia during and after COVID-19 infection, particularly among vulnerable groups and hospitalized patients [8, 9]. Sleep problems are of particular concern because they may persist after recovery and can negatively affect daily functioning, quality of life, and overall mental health. In addition, insomnia may coexist with depressive symptoms, and this relationship may be especially relevant in patients recovering from severe infectious illness.

Given the prolonged course of the COVID-19 pandemic and the growing recognition of its post-acute neuropsychiatric sequelae, evaluating insomnia in patients who have recovered from COVID-19 is clinically important. Patients who required hospitalization may represent a higher-risk group because of greater disease severity, treatment-related stress, and post-discharge psychological burden. However, local evidence on the prevalence of insomnia after hospitalization for COVID-19 and its association with depression remains limited. Although a few local studies have investigated this issue in the general population [10, 11], most available research has focused on specific groups, such as medical students and healthcare workers [12–14]. Therefore, the present study aimed to determine the prevalence of insomnia and its association with depression among patients who had recovered from COVID-19 after hospitalization at Shahid Beheshti Hospital in Zanjan in 2021.

## 2 Methods

### 2.1 Study design and setting

This analytical cross-sectional study was conducted to determine the prevalence of insomnia among patients who had recovered from COVID-19 and to assess its association with depressive symptoms. The study was performed at Shahid Beheshti Hospital, affiliated with Zanjan University of Medical Sciences, Zanjan, Iran, in 2023. Interviews were performed between Februery to May 2023.

### 2.2 Data

The study population consisted of all patients who were hospitalized at Shahid Beheshti Hospital between 13 April and 5 May 2021 and between 31 August and 10 September 2021 because of COVID-19. Sampling was performed using a census method; therefore, all eligible patients were considered for inclusion and no formal sample size calculation was performed. A total of 70 recovered patients were included in the final analysis.

Patients were eligible for inclusion if they: 1) agreed to participate in the study; 2) were aged 15–50 years; 3) had a previous diagnosis of COVID-19 confirmed by polymerase chain reaction (PCR) test; and 4) had recovered from COVID-19 for at least 6 months at the time of assessment. Patients were excluded if they had: 1) a prior history of psychiatric disorders, including depression and sleep disorders, based on DSM-5 criteria; 2) substance or psychotropic drug dependence, with the exception of nicotine and caffeine; or 3) unwillingness to continue participation.

After obtaining the required approvals, the researchers referred to the medical records unit of Shahid Beheshti Hospital and identified all patients who had been hospitalized because of COVID-19 during the study period. Demographic and clinical information was first extracted from medical records using a structured checklist. Subsequently, the participants were contacted by telephone using the contact numbers recorded in their hospital files. During the telephone interview, data on depressive symptoms and insomnia severity were collected using standardized questionnaires.

Data collection included demographic characteristics such as age, sex, marital status, educational level, employment status, place of residence, monthly income, and history of non-psychiatric underlying disease.

**Table 1:** Description and operational definitions of study variables.

| Variable Name | Quant. | Qual. | Operational Definition |
| --- | --- | --- | --- |
| Insomnia | ✓ |  | Score obtained on ISI |
| Depressive Symptoms | ✓ |  | Score obtained on BDI-II |
| Gender |  | ✓ | Male / Female |
| Age | ✓ |  | Age in years |
| Marital Status |  | ✓ | Single / Married |
| Residence |  | ✓ | Urban / Rural |
| Non-psychiatric Comorbidity |  | ✓ | Chronic illness or physical disability existence |
| Education Level |  | ✓ | Academic / Non-academic |
| Monthly Income |  | ✓ | Below 50M / 50–100M / Above 100M IRR |

#### Insomnia Severity Index

Insomnia severity was assessed using the Insomnia Severity Index (ISI) [17], a self-report instrument developed by Morin. The ISI consists of seven items that evaluate difficulties with sleep onset, sleep maintenance, early morning awakening, satisfaction with the current sleep pattern, interference of sleep problems with daily functioning, noticeability of impairment attributed to sleep difficulties, and the degree of distress or concern caused by sleep problems. Each item is rated on a 0–4 scale, yielding a total score of 0 to 22, with higher scores indicating greater insomnia severity. The standard scoring categories are as follows: 0–7, no clinically significant insomnia; 8–14, subthreshold insomnia; 15–21, moderate clinical insomnia; and 22–28, severe clinical insomnia. The validity and reliability of the Persian version of the ISI have previously been confirmed in Iran [18].

#### Beck Depression Inventory-II

Depressive symptoms were assessed using the Beck Depression Inventory-II (BDI-II) [15]. This self-report instrument consists of 21 items assessing emotional, cognitive, motivational, and somatic symptoms of depression. Each item is scored on a scale from 0 to 3, yielding a total score ranging from 0 to 63. Based on standard scoring, depression severity was categorized as follows: 0–9, minimal or no depression; 10–13, mild depression; 14–20, moderate depression; and 21–27, severe depression; and above 27 very severe depression. The Persian version of the questionnaire has previously demonstrated acceptable validity and reliability [16].

### 2.3 Statistical analysis

Descriptive statistics were used to summarize the data. Categorical variables were presented as frequency and percentage, whereas continuous variables were expressed as mean **±** standard deviation, with minimum and maximum values where appropriate.

The normality of quantitative continuous variables was assessed using the Kolmogorov-Smirnov test. Depending on the distribution of the variables, comparisons between groups were performed using the independent-samples t test or one-way analysis of variance (ANOVA) for normally distributed variables, and the Mann-Whitney U test or Kruskal-Wallis test for non-normally distributed variables. Categorical variables were compared using the chi-square test or Fisher’s exact test, as appropriate.

To identify factors associated with insomnia severity, univariable linear regression analysis was first performed. Variables with a p-value *<* 0.20 in univariable analysis were entered into a multivariable linear regression model to adjust for potential confounding factors. *β* coefficients and 95% confidence intervals (CIs) were reported for both crude and adjusted models. A p-value *<* 0.05 was considered statistically significant.

### 2.4 Ethical considerations

This study was approved by the Ethics Committee of Zanjan University of Medical Sciences, Zanjan, Iran, under the ethics code IR.ZUMS.REC.1401.194 on 18 September 2022. The objectives and procedures of the study were explained to all participants, and informed consent was obtained. Participant confidentiality was maintained throughout the study, and no additional costs were imposed on the patients. The study was conducted in accordance with the ethical principles of the Declaration of Helsinki [19].

## 3 Results

A total of 70 patients who had recovered from COVID-19 after hospitalization at Shahid Beheshti Hospital were included in the analysis. The mean age of the participants was 36.75± 8.1 years. Overall, 43 participants (61.4%) were women and 47 (67.1%) were married. 47 participants (67.1%) were employed, and 50 (71.4%) had an academic educational level. Most participants lived in urban areas (68 patients, 97.1%). Regarding monthly income, the most common category was 50 to 100 million IRR observed in 37 participants (52.9%). A history of non-psychiatric underlying disease was reported by 11 participants (15.7%). The demographic and clinical characteristics of the study population are summarized in table 2.

**Table 2:**
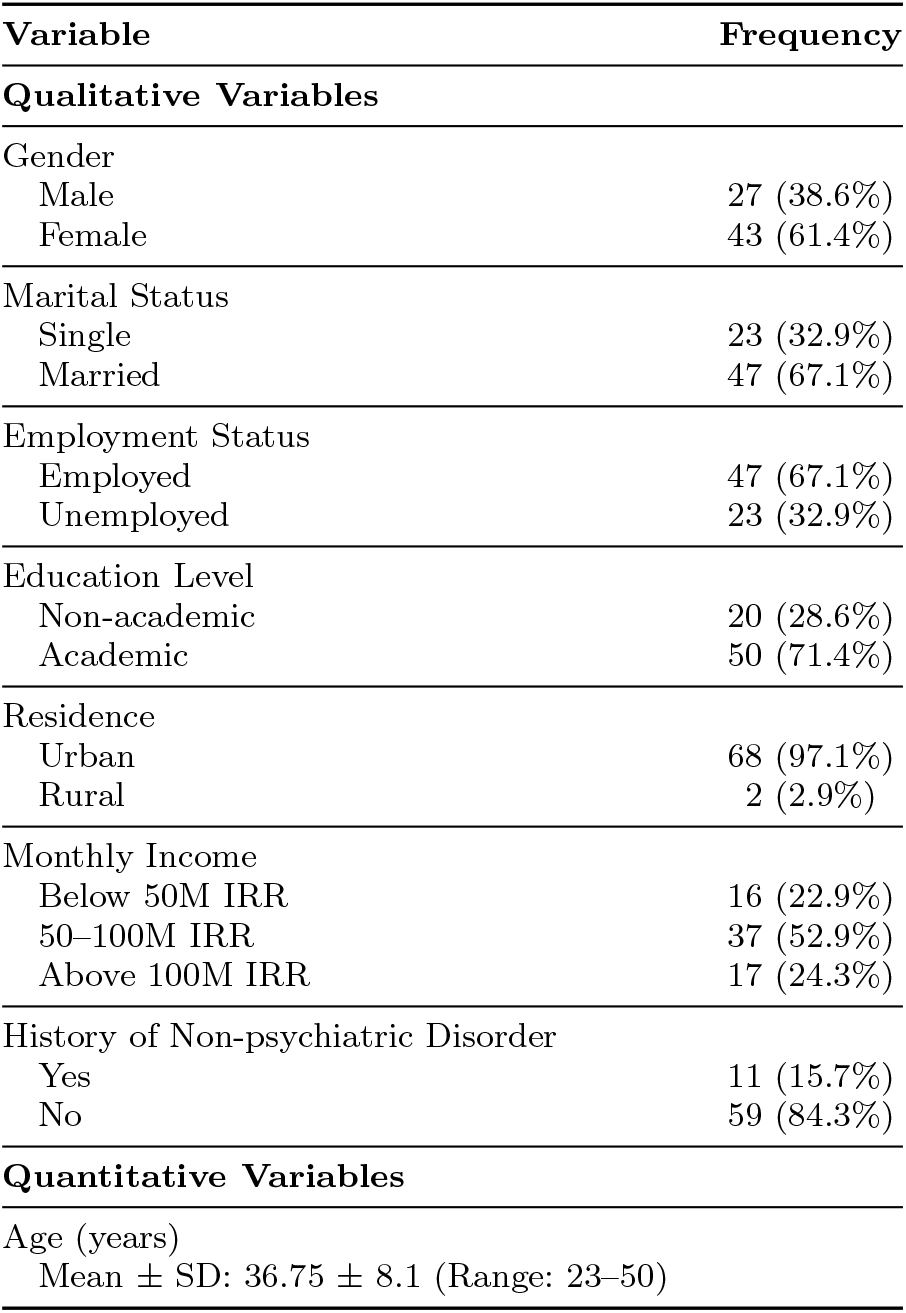
Demographic Characteristics of the Study Participants (n = 70)

The mean Beck Depression Inventory-II score was 8.63 ± 7.94. Based on the depression severity categories, 49 participants (70.0%) had no depression. Mild and moderate depressive symptoms were each observed in 7 participants (10.0%), whereas severe depressive symptoms in 3 participants (4.3%), and very severe depressive symptoms in 4 participants (5.7%). The distribution of depressive symptom severity is presented in figure 1.

**Fig. 1:**
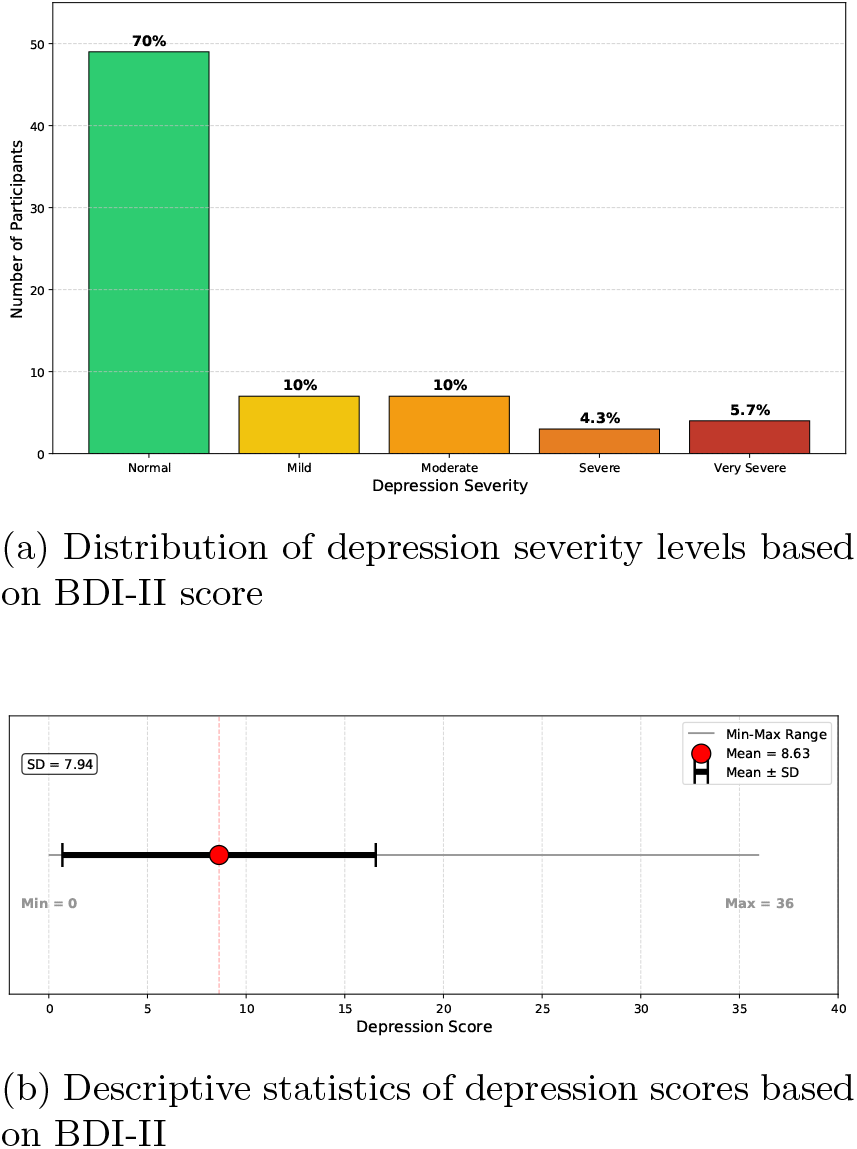
Depression severity and descriptive statistics in the study population (n = 70). (a) Distribution of participants across depression severity categories. (b) Dot plot showing mean (8.63), standard deviation (7.94), and range (0–36) of depression scores.

The mean Insomnia Severity Index score was 8.06± 6.57. 42 participants (60.0%) had no clinically significant insomnia. Subthreshold insomnia was observed in 16 participants (22.9%), moderate clinical insomnia in 9 participants (12.9%), and severe clinical insomnia in 3 participants (4.3%). Therefore, 28 participants (40.0%) had at least subthreshold insomnia, and 12 participants (17.1%) met the ISI criteria for moderate or severe clinical insomnia. The distribution of insomnia severity is shown in figure 2.

**Fig. 2:**
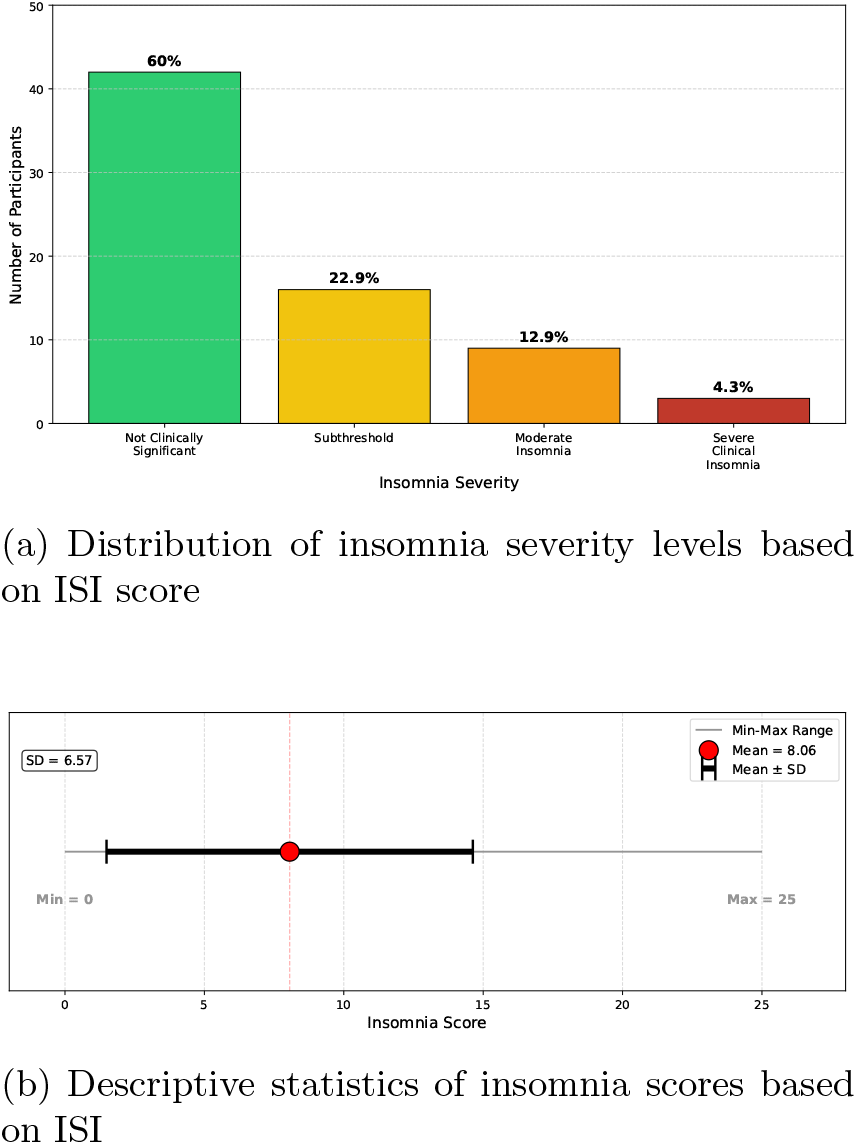
Insomnia severity and descriptive statistics in the study population (n = 70). (a) Distribution of participants across insomnia severity categories. (b) Dot plot showing mean (8.06), standard deviation (6.57), and range (0–25) of insomnia scores.

In univariable linear regression analysis, depressive symptom score and educational level were associated with insomnia severity at the pre-defined significance level for entry into the multivariable model. Each one-point increase in BDI-II score was associated with a 0.52-point increase in ISI score (*β* = 0.52; 95% CI: 0.36 to 0.67; *p <* 0.001). Academic education was associated with a lower insomnia severity score compared with non-academic education (*β* = -3.52; 95% CI: -6.91 to -0.13; p = 0.042). Other demographic and clinical variables, including age, sex, marital status, employment status, place of residence, monthly income, and history of non-psychiatric underlying disease, were not significantly associated with insomnia severity in the univariable analysis.

In the multivariable linear regression model, after adjustment for potential confounding variables, depressive symptom score remained independently associated with insomnia severity. Each one-point increase in BDI-II score was associated with a 0.50-point increase in ISI score (adjusted *β* = 0.50; 95% CI: 0.34 to 0.66; *p <* 0.001).

After adjustment, educational level was no longer identified as an independent predictor of insomnia severity. The univariable and multivariable regression results are shown in table 3.

**Table 3:** Predictors of Insomnia in COVID-19 Survivors: Univariate and Multivariable Linear Regression Models.

| Variable | $\beta$ | Std. Error | 95% CI | P-Value |
| --- | --- | --- | --- | --- |
| **Univariable regression model** |  |  |  |  |
| Depression score (BDI-II) | 0.52 | 0.08 | [0.36, 0.67] | <001* |
| Age (years) | 0.03 | 0.09 | [-0.23, 0.16] | 0.753 |
| Gender (*ref. Male*) |  |  |  |  |
| Female | -0.40 | 1.62 | [-3.65, 2.84] | 0.805 |
| Marital status (*ref. Single*) |  |  |  |  |
| Married | -0.13 | 1.68 | [-3.49, 3.23] | 0.938 |
| Employment (*ref. Employed*) |  |  |  |  |
| Unemployed | -0.64 | 1.68 | [-4.00, 2.71] | 0.703 |
| Education (*ref. Non-academic*) |  |  |  |  |
| Academic | -3.52 | 1.70 | [-6.91, 0.13] | 0.042* |
| Residence (*ref. Urban*) |  |  |  |  |
| Rural | 0.43 | 4.75 | [-9.05, 9.90] | 0.929 |
| Monthly income (*ref. < 50M IRR*) |  |  |  |  |
| 50–100M IRR | -2.49 | 1.97 | [-6.42, 1.43] | 0.210 |
| > 100M IRR | -2.71 | 2.29 | [-7.28, 1.86] | 0.241 |
| Non-psychiatric illness (*ref. No*) |  |  |  |  |
| Yes | 2.70 | 2.15 | [-1.59, 6.99] | 0.213 |
| **Multivariable regression model** |  |  |  |  |
| Depression score | 0.50 | 0.08 | [0.34, 0.66] | <001** |
| Education (*ref. Non-academic*) |  |  |  |  |
| Academic | -1.18 | 1.42 | [-4.01, -1.64] | 0.406 |

Notes: CI = confidence interval; IRR = Iranian rial; M = million; *ref.* = reference category. Asterisks indicate statistically significant associations: \* significant in the univariable regression model; \*\* significant in the multivariable regression model.

Overall, the findings showed that depressive symptoms and insomnia symptoms were relatively common among patients who had recovered from COVID-19 after hospitalization. In addition, depressive symptom severity was independently associated with greater insomnia severity in this study population.

## 4 Discussion

In this analytical cross-sectional study of patients who had recovered from COVID-19 after hospitalization, insomnia symptoms were common and were closely associated with depressive symptom severity. Overall, 40.0% of participants had at least subthreshold insomnia according to the Insomnia Severity Index, and 17.1% had moderate or severe clinical insomnia. Depressive symptoms were also frequent, with 30.0% of participants showing some degree of depression based on the Beck Depression Inventory-II. In regression analysis, depressive symptom score was the only factor that remained independently associated with insomnia severity after adjustment for potential confounders. These findings suggest that sleep disturbance and depressive symptoms may persist after the acute phase of COVID-19, particularly among patients who required hospitalization.

The observed prevalence of insomnia in the present study is consistent with the growing evidence that sleep problems are among the common neuropsychiatric and post-acute consequences of COVID-19, although the exact prevalence varies according to study population, time since infection, disease severity, assessment instrument, and sociocultural context [8, 9].

Several mechanisms may explain the persistence of insomnia after COVID-19. Hospitalized patients may experience fear of death, uncertainty about prognosis, isolation from family members, disruption of daily routines, and exposure to stressful hospital environments, all of which may contribute to sleep disturbance [20–22]. In addition, biological mechanisms such as systemic inflammation, immune dysregulation, autonomic activation, hypoxia, and possible neuroinflammatory effects of SARS-CoV-2 have been proposed as contributors to post-COVID neuropsychiatric symptoms, including insomnia [23–25]. Post-discharge symptoms such as fatigue, dyspnea, pain, reduced physical activity, and concerns about reinfection or long-term disability may also maintain sleep problems after apparent recovery [26].

Depressive symptoms were also relatively frequent in this sample. This finding is in line with previous studies reporting increased rates of depression among patients after COVID-19 infection, particularly among those who experienced severe disease, hospitalization, or prolonged recovery [8, 9]. Depression after COVID-19 may be related not only to the psychological burden of infection and hospitalization, but also to social isolation, financial stress, reduced occupational functioning, and persistent physical symptoms [20, 23, 26]. However, because depression was assessed using the BDI-II, the findings should be interpreted as depressive symptom severity rather than a confirmed clinical diagnosis of major depressive disorder. One important finding of the present study was the independent association between depressive symptom severity and insomnia severity. In the adjusted linear regression model, each one-point increase in BDI-II score was associated with an approximately 0.50-point increase in ISI score. This association is clinically plausible, as insomnia and depression are strongly interconnected conditions. Insomnia may increase vulnerability to depression through emotional dysregulation, impaired stress tolerance, cognitive hyperarousal, and disruption of circadian rhythms. Conversely, depression may contribute to insomnia through rumination, early morning awakening, reduced daytime activity, and changes in neuroendocrine and inflammatory pathways [27]. Therefore, in patients recovering from COVID-19, insomnia and depressive symptoms may reinforce each other and contribute to persistent impairment in quality of life and daily functioning.

In the univariable analysis, educational level was associated with insomnia severity, with lower insomnia scores observed among participants with academic education. However, this association did not remain an independent predictor after adjustment. This finding suggests that the apparent relationship between education and insomnia may have been influenced by other factors, particularly depressive symptom severity. Previous studies have reported mixed findings regarding the relationship between education and sleep outcomes during the COVID-19 pandemic [28, 29]. Educational level may affect access to health information, coping strategies, health literacy, socioeconomic stability, and help-seeking behavior, but its independent role in post-COVID insomnia requires further investigation. Other demographic and clinical variables, including age, sex, marital status, employment status, place of residence, income level, and history of non-psychiatric underlying disease, were not significantly associated with insomnia severity in this study. This may indicate that depressive symptom burden had a stronger relationship with insomnia than the measured demographic characteristics in this sample. However, the absence of significant associations should be interpreted cautiously because the study included only 70 participants, which limits statistical power to detect small or moderate effects. In addition, some categories included few participants, such as rural residence, which may have reduced the ability to identify meaningful sub-group differences.

The findings have important clinical implications. Due to their high prevalence, screening for insomnia and depressive symptoms should be considered in follow-up care for patients discharged after hospitalization for COVID-19. Early identification of sleep problems may help prevent chronic insomnia and reduce the risk of associated psychiatric morbidity. Integrated post-COVID care models should include mental health assessment, psychoeducation about sleep hygiene, and referral pathways for psychological or psychiatric intervention when needed. Evidence-based interventions such as cognitive behavioral therapy for insomnia, stress-management strategies, and treatment of clinically significant depression may be beneficial in this population [30, 31].

This study has several strengths. It focused on recovered patients who had been hospitalized for COVID-19, a group that may be at increased risk for post-acute psychological complications. The study also used standardized instruments, including the BDI-II and ISI, both of which have previously been used in Iranian population and have acceptable psychometric properties [15, 17]. Furthermore, the analysis examined the association between depressive symptoms and insomnia while considering demographic and clinical variables as potential confounders.

Several limitations should also be acknowledged. First, the cross-sectional design prevents any causal interpretation of the relationship between depressive symptoms and insomnia. It is not possible to determine whether depressive symptoms contributed to insomnia, insomnia contributed to depressive symptoms, or both developed as related consequences of COVID-19 and hospitalization. Second, the sample size was relatively small and was drawn from a single hospital, which may limit the generalizability of the findings to other settings or populations. Third, insomnia and depressive symptoms were assessed by self-report questionnaires through telephone interviews, which may be subject to recall bias, reporting bias, and differences in participant understanding of questionnaire items. Another limitation is the lack of detailed information on COVID-19 severity and treatment-related factors. Variables such as duration of hospitalization, intensive care unit admission, and persistence of physical symptoms after discharge were not considered. These factors may be relevant to both insomnia and depression and should be considered in future studies.

Future research should include larger, multicenter studies with longitudinal designs to clarify the temporal relationship between COVID-19, insomnia, and depressive symptoms. Repeated assessments after discharge could help determine whether sleep disturbance improves over time or persists as part of a post-COVID syndrome. Future studies should also include clinical diagnostic interviews, more detailed clinical data regarding acute COVID-19 severity and post-discharge physical symptoms. Such evidence would help identify high-risk patients and inform targeted interventions.

## 5 Conclusion

In conclusion, insomnia and depressive symptoms were relatively common among patients who had recovered from COVID-19 after hospitalization. Depressive symptom severity was independently associated with greater insomnia severity, suggesting that psychological distress may play an important role in post-COVID sleep disturbance. These findings support the need for routine screening and early mental health interventions in patients recovering from hospitalized COVID-19, with particular attention to the coexistence of insomnia and depressive symptoms.

## Data Availability

Data produced in the present study are not available to public

## Acknowledgement

The authors acknowledge the use of GPT-5.5, an artificial intelligence language model, for assistance with language editing, improving readability, and drafting the manuscript. The authors reviewed, revised, and verified all AI-assisted content and take full responsibility for the accuracy, integrity, and final content of the manuscript.

